# Unraveling the Molecular Landscape of Congenital Pseudoarthrosis of the Tibia: Insights from a Comprehensive Analysis of 162 Probands

**DOI:** 10.1101/2023.06.28.23292011

**Authors:** Guanghui Zhu, Nan Li, Yu Zheng, Shunyao Wang, Ge Yang, Yaoxi Liu, Zongren Xu, Hui Huang, Huanhuan Peng, Haibo Mei

## Abstract

Congenital pseudarthrosis of the tibia (CPT, HP:0009736), commonly known as bowing of the tibia, is a rare congenital tibia malformation characterized by spontaneous tibial fractures and the difficulty of reunion after tibial fractures during early childhood, with a very low prevalence between 1/250,000∼1/140,000. While 80%–84% of CPT cases present with neurofibromatosis type 1, caused by the mutations in *NF1*, the underlying cause of CPT is still unclear. Considering the congenital nature and the low prevalence of CPT, we hypothesized that the rare genomic mutations may contribute to CPT. In this study, we conducted whole exome sequencing on 159 patients with CPT and full-length transcriptome sequencing on an additional 3 patients with CPT. The data analysis showed there were 179 significantly up-regulated genes which were enriched in 40 biological processes among which 21 biological processes hold their loss of function (LoF) excesses between 159 cases against 208 controls from 1000 Genomes Project. From those 21 biological processes with LoF excesses, there were 259 LoF-carried genes among which 40 genes with 56 LoF variations in 63 patients were enriched in osteoclast differentiation pathway (hsa04380) with its 3 directly regulated pathways including MAPK signaling pathway (hsa04010), calcium signaling pathway (hsa04020) and PI3K-Akt signaling pathway (hsa04151), as well as fluid shear stress and atherosclerosis pathway (hsa05418) while 12 patients carried 9 LoF variations in the *NF1* gene. The rare LoF variations in these pathways accounted for ∼39.6% of this CPT cohort. These findings shed light on the novel genetic mutations and molecular pathways involved in CPT, providing a new framework for understanding how the genetic variations regulate the biological processes in the pathology of CPT and indicating potential next directions to further elucidate the pathogenesis of CPT.

## Introduction

Congenital pseudarthrosis of the tibia (CPT) is a rare orthopedic disease with an incidence between 1 in 140,000 and 1 in 250,000 live births [1]. It is characterized by the pseudarthrosis or pathological fractures of the anterolateral part of the tibia in early life, resulting in bowing, narrowing of the medullary canal, or the presence of a cyst. This condition poses a significant surgical challenge due to recurrent fractures and the inability to achieve bone union [2]. Congenital anterolateral bowing of the tibia is generally considered a precursor of CPT and is commonly associated with neurofibromatosis type 1(NF1), a common autosomal dominant genetic disorder [3]. Previous research has reported that 80%–84% of CPT cases present with NF1(NF1-CPT) in the epidemiology and the genomic mutations in the *NF1* gene have been linked to CPT in the genomics [4–7]. The double inactivation model has been proposed as a hypothesis to explain these associations [8–11]. However, other researchers have suggested that the molecular pathogenesis of NF1-CPT may not be entirely explained by second mutations or loss of heterozygosity of the *NF1* gene [7, 12, 13].

Recent investigations have shed light on the molecular basis of CPT, for example, *NF1* gene has been reported to be involved in bone remodeling and mineralization [14]. Inhibited bone formation and stimulated bone resorption have also been reported in patients with CPT who harbor *NF1* mutations [15]. Furthermore, the involvement of FGFR3 signaling in cartilage-to-bone transformation for bone repair [16] and the contribution of vasoactive intestinal peptide through ERK and NF-κB signal pathways have been implicated in CPT [17]. Differentially expressed proteins (DEPs) found in serum-derived exosomes of CPT patients have been shown to inhibit bone formation and stimulate bone resorption[18] while DEPs from in the tibia periosteum tissues are mainly involved in cell matrix assembly, cell adhesion, AKT-PI3K signal pathway activation, and vascular agglutination of CPT patients [19]. These conclusions indicate the intricate biological processes and molecular regulation pathways associated with bone homeostasis potentially contribute to CPT [20–23]. However, a comprehensive understanding of the etiology of CPT is still elusive due to the rarity of CPT and limited samples in previous studies.

To address this gap, we employed an approach used to investigate the severe pediatric developmental disorders with a low prevalence and the complex genetic architecture, aiming to identify the low-frequency gene mutations in 159 sporadic CPT patients. This approach have been successful in elucidating the genetic basis of other neurodevelopmental disorders, including epilepsy [24–28], craniosynostosis [29–31], autism [32–40] and congenital heart disease [41–46]. Specially the rare protein-damaging variations (PDV), particularly loss of function (LoF) mutations, have been identified a main contributor to congenital developmental diseases characterized by low prevalence and early onset. In light of this, we hypothesized that CPT can also be explained, at least partially, by the rare LoF variants with incomplete penetrance. To test this hypothesis, we conducted whole exome sequencing (WES) on a substantial cohort of 159 individuals affected by idiopathic CPT. Additionally, we performed a full-length transcriptome sequencing on 3 CPT patients to validate the reported biological processes and detect the new ones associated with CPT.

In this study, we present our comprehensive genetic analysis, aiming to uncover the complex genetic architecture and molecular mechanism underlying CPT. Our findings have the potential to enhance the understanding of CPT pathogenesis and provide valuable guidance for the development of novel therapeutic approaches targeting the identified genetic factors.

## Results

To comprehensively explore the genetic basis and biological processes underlying CPT, a cohort of 159 along with an additional 3 Chinese Han CPT patients, with an average age of 42 months, were enrolled for WES and full-length transcriptome sequencing respectively. In the WES, targeted bases in each sample were sequenced a mean of 136 times by independent reads, with 98.73% reading 20 or more times. Following variation calling, a totally of 286,945 single nucleotide polymorphisms (SNPs) and 27,807 insertions/deletions (InDels) were detected. For the transcriptome sequencing, an average of 4,816,579 reads with a length of 787 bp were obtained for each sample.

### 40 bone homeostasis related biological processes were up-regulated in CPT patients

To get insights into the CPT-related pathways, we performed nanopore sequencing on the periosteum tissues from three CPT patients against two healthy individuals. The differential expression analysis of transcriptome identified 179 genes up-regulated and 134 down-regulated genes excluding *NF1* gene (|logFC| > 2, FDR < 0.01) in patients with CPT (Fig1 a and b). Functional enrichment analysis of those up-regulated genes uncovered 40 significantly enriched biological processes (threshold P=0.01). These enriched biological processes encompassed skeletal system development (logP=-11.96), tissue remodeling including bone resorption, cell matrix assembly (logP=-10.37) [47, 48], regulation of cell adhesion (logP=-8.44), regulation of MAPK cascade [49] (logP=-5.22) and autophagy in bone metabolism [50, 51], which have been associated with bone formation and bone remodeling [19, 52–54]. Additional enriched biological progress included ossification (e.g., bone mineralization and endochondral ossification) [55], myeloid leukocyte differentiation (e.g., osteoclast differentiation and osteoclast development) [56, 57], epithelial cell proliferation (e.g., angiogenesis) [58–61], inorganic ion homeostasis [62, 63], regulation of hormone levels [64, 65], negative regulation of response to external stimulus [57, 66], Vitamin D receptor pathway [67–69], cell migration and chemotaxis [70] (Fig.1.c and supplementary table 1, threshold P= 0.05/100=5e-4 by Bonferroni correction). Some of these processes have been reported in the DEPs of CPT patients [18, 19, 53] and also known to play crucial important role in the metabolism of bone homeostasis. The enriched biological processes identified in the transcriptome analysis provide a comprehensive reflection of the etiology of CPT.

**Fig1.**
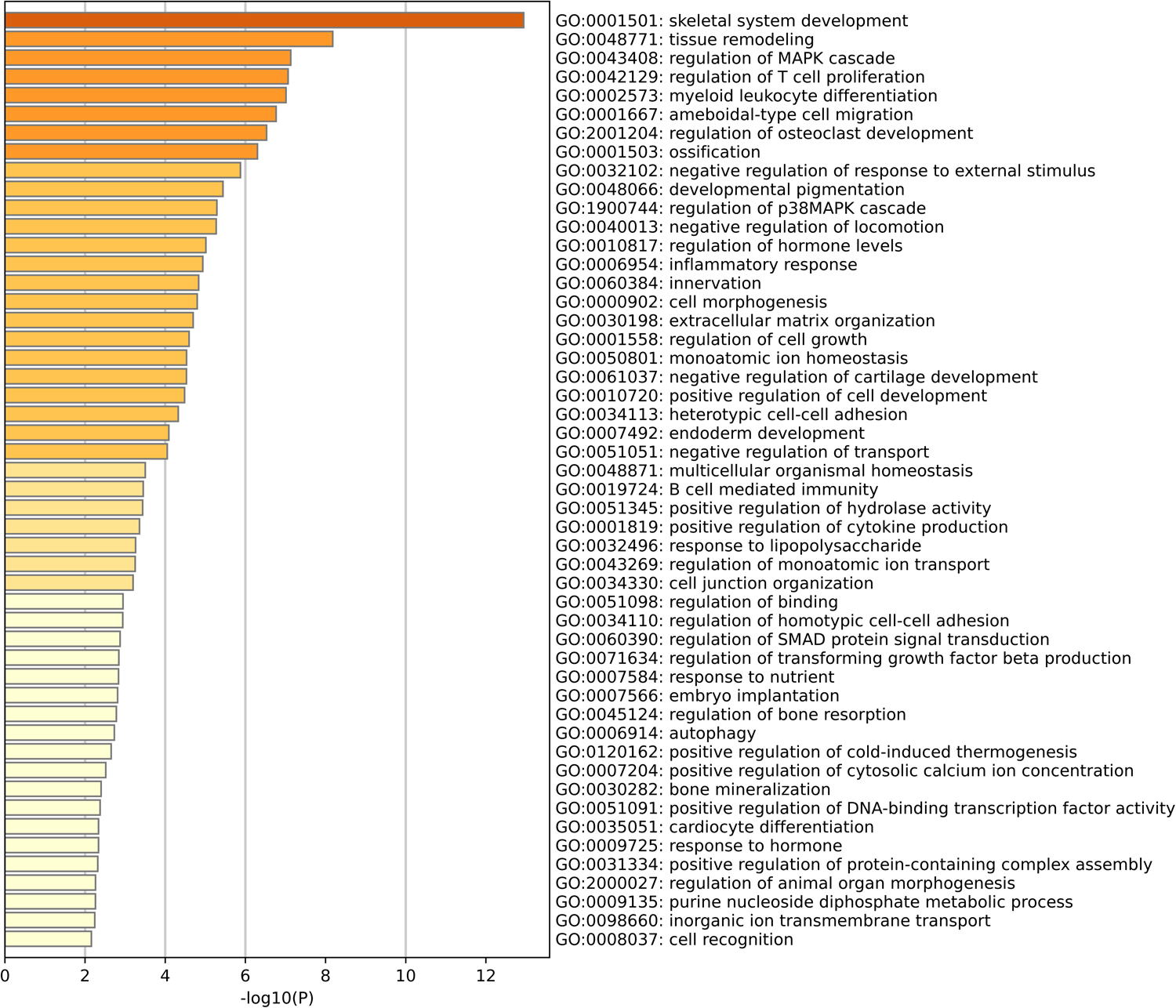
Identification of differentially expressed genes associated with CPT in the transcriptome sequencing (a) Volcano plot for the comparison between the 3 CPT (JG159_nonNF1, JG250_NF1 and JG252_NF1) and 2 healthy individuals (GDZ001_Control and GDZ002_Control). The cutoff values fold change >2 and FDR < 0.01 were utilized to identify differentially expressed genes. Non-changed genes were shown in black color. Red color is indicative of the up-regulated genes and the down-regulated genes. (b) Heatmap plot of differentially expressed genes. The Pearson correlation distance metric and the average linkage clustering algorithm were used. (c) the GO enrichment of the 179 up-regulated genes in the transcriptome. X-axis indicates the -log(P) in which P is the probability that the genes are enriched in the biological process in GO database. Here the threshold of -log(P) is set as 2, indicating the P value 0.01.

### 21 of 40 biological processes beared the LoF excesses in CPT patients

Considering the rare prevalence and early onset of CPT, we postulate that some or all of the 40 biological processes may bear a significant burden of rare LoF mutations if they contribute to CPT pathogenesis. To test this hypothesis, we analyzed the genetic burden of LoF variations from WES data. After variation calling and filtering, 1,174 confidant LoF single nucleotide variants (SNVs) in 1,037 genes (an average of 7.38 mutations per case with a mutation rate of 4.34e-9) were selected for the burden test between 159 CPT cases and 208 healthy Chinese Han controls from the 1000 Genomes Project. Among all the 40 significantly enriched and up-regulated biological processes in transcriptome, 21 processes showed LoF excesses under the threshold P <0.01 and OR > 10 (Fig2, Table 1), indicating their potential relevance to CPT. Additionally, the burden test of rare LoF mutations showed no significant excess of LoF (Bonferroni corrected threshold P-value 0.05/1,217=4.1e-5) for each gene except for *NF1* (p= 5.61357e-05) which was reported in the previous studies [4, 7]. This result suggests that the 21 CPT-related candidate biological processes, which exhibit a significant rare LoF excess, cannot be detected by the burden test for single gene. Furthermore, the other 19 biological processes significantly up-regulated in the transcriptome and enriched in the Gene Ontology (GO) database did not show a rare LoF excess, indicating that rare LoF mutations alone cannot entirely explain the pathogenicity and the complex genetic architecture of CPT.

**Fig2.**
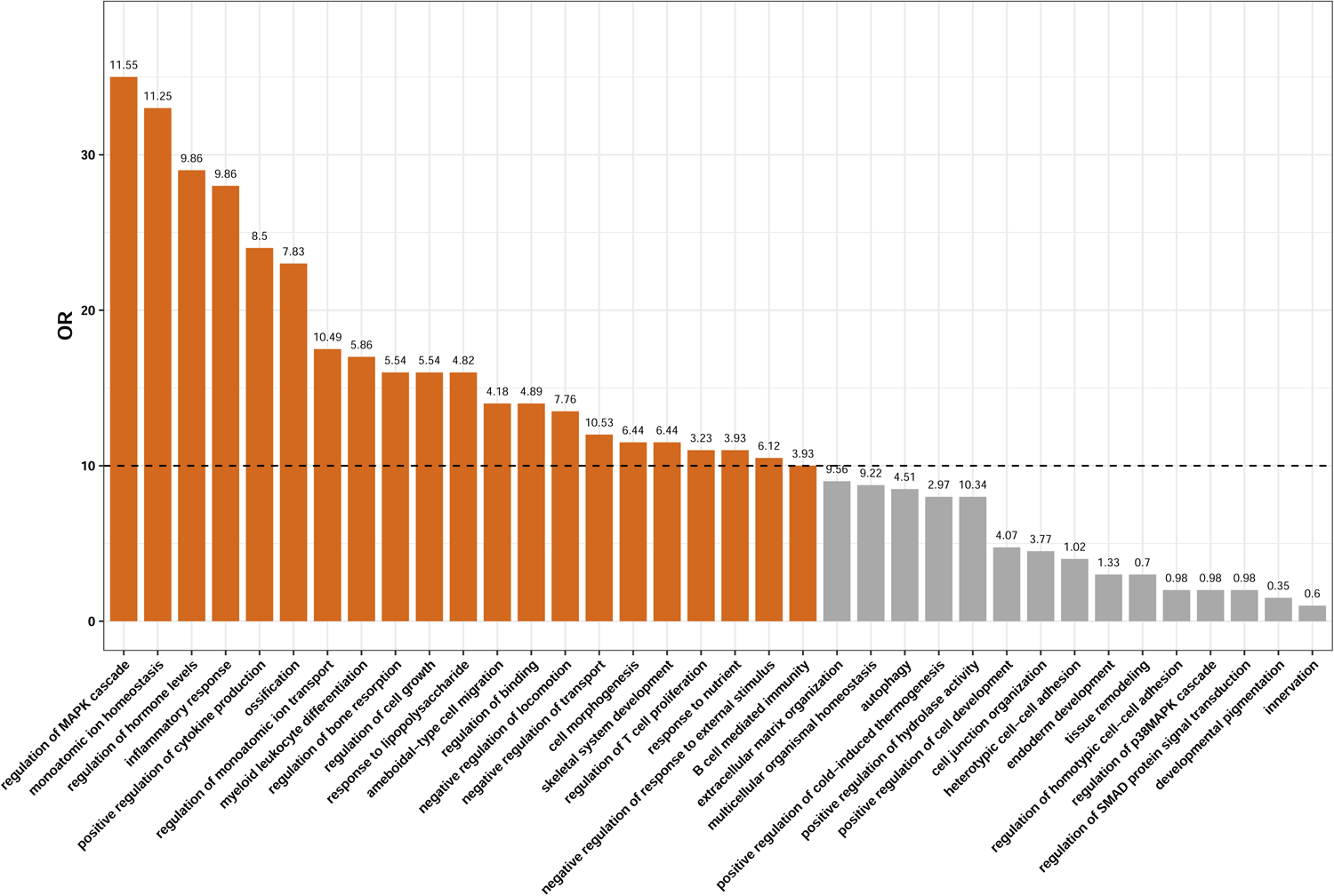
The 179 up-regulated genes in patients with CPT significantly enriched in 40 biological processes in the GO biological processes (the Bonferroni corrected P= 0.05/100=5e-4) among which 21 revealed the LoF excesses (under the criteria OR >10 and P > 0.01). X-axis indicates the biological processes from GO database and Y-axis indicates the odd ratio (OR) that the LoF mutations enrich in the biological process between 159 CPT cases and 208 controls. The number above the bar in each process is the P values indicating the enrichment of LoF mutations in this process.

**Table 1.**
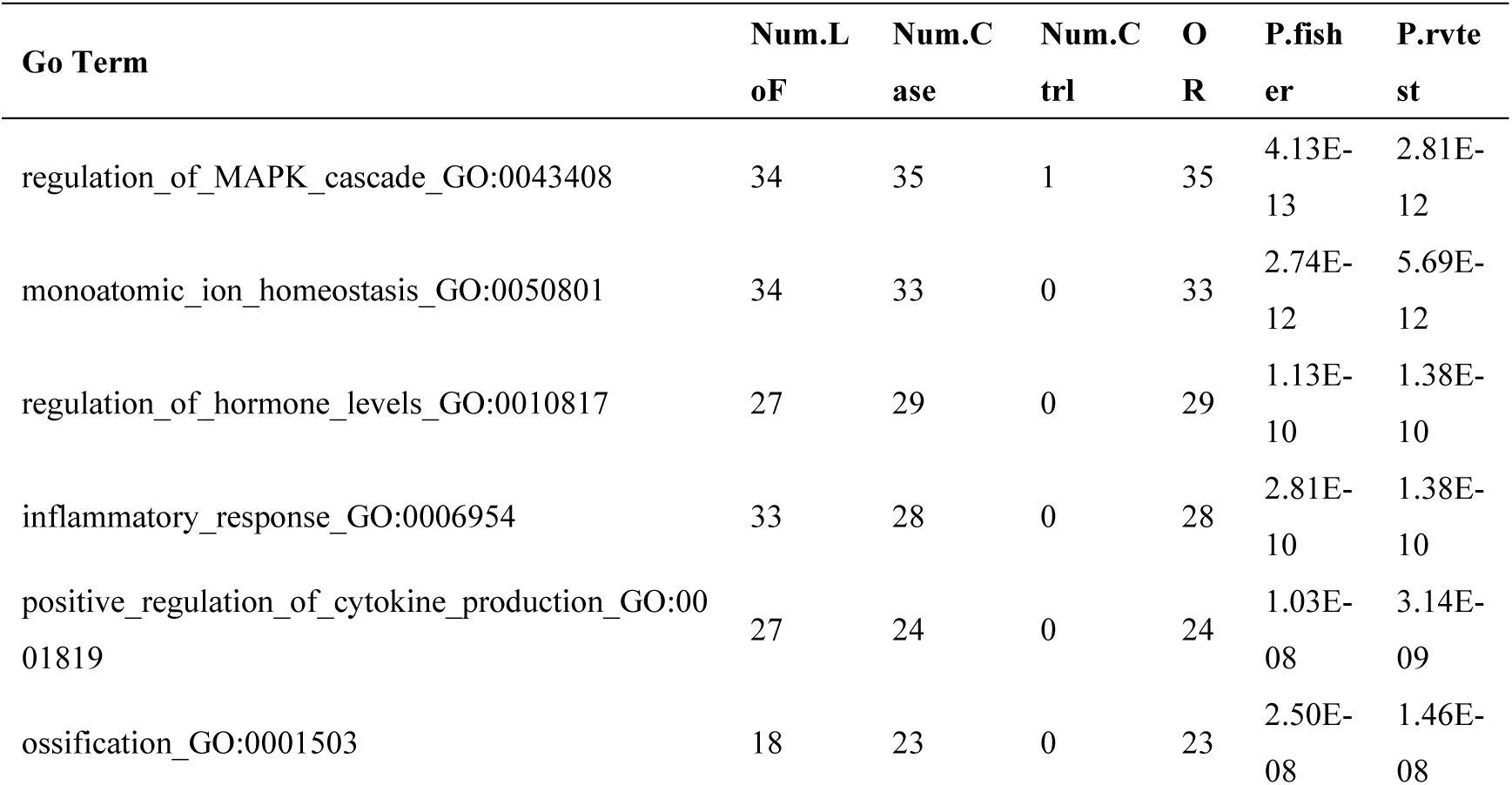
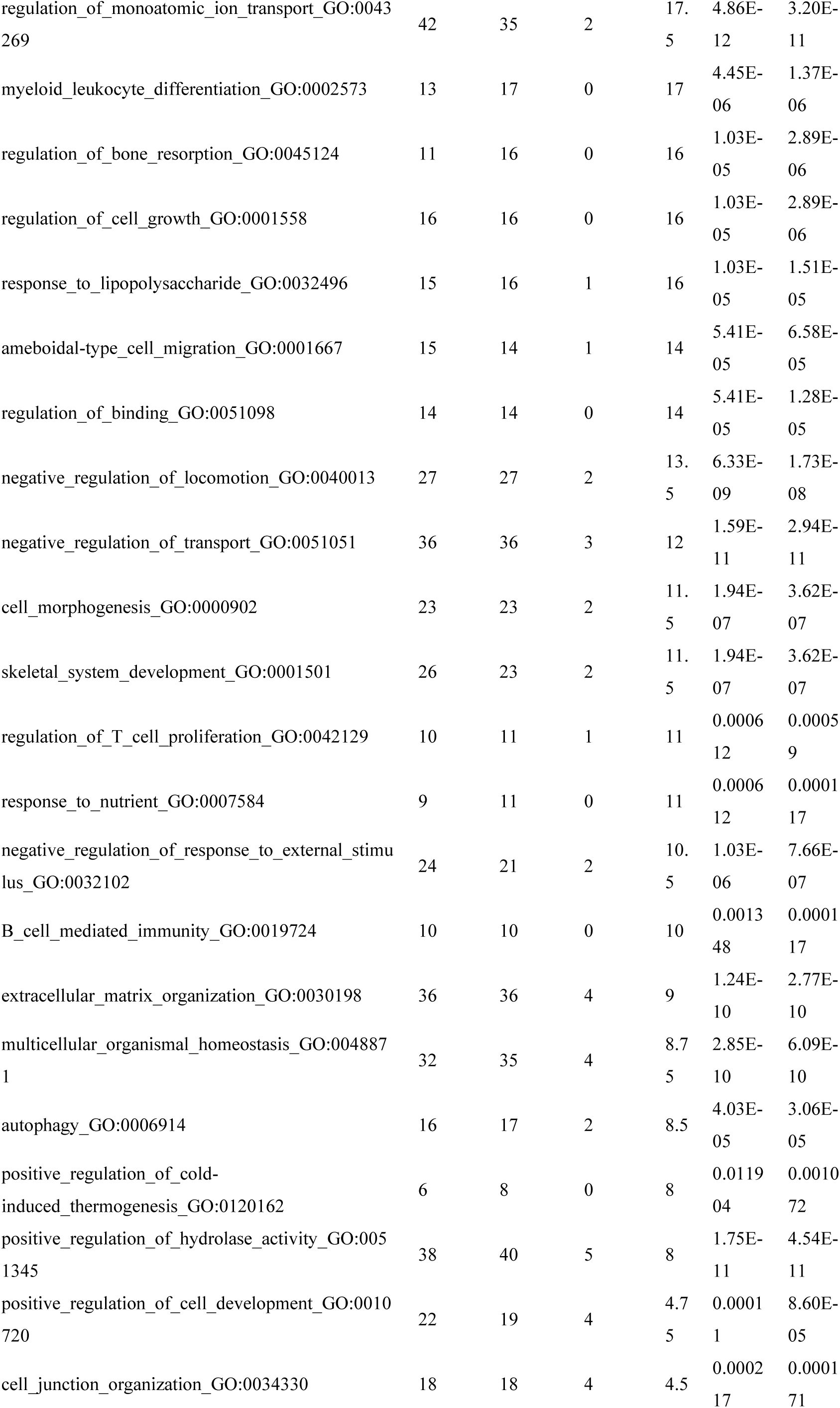
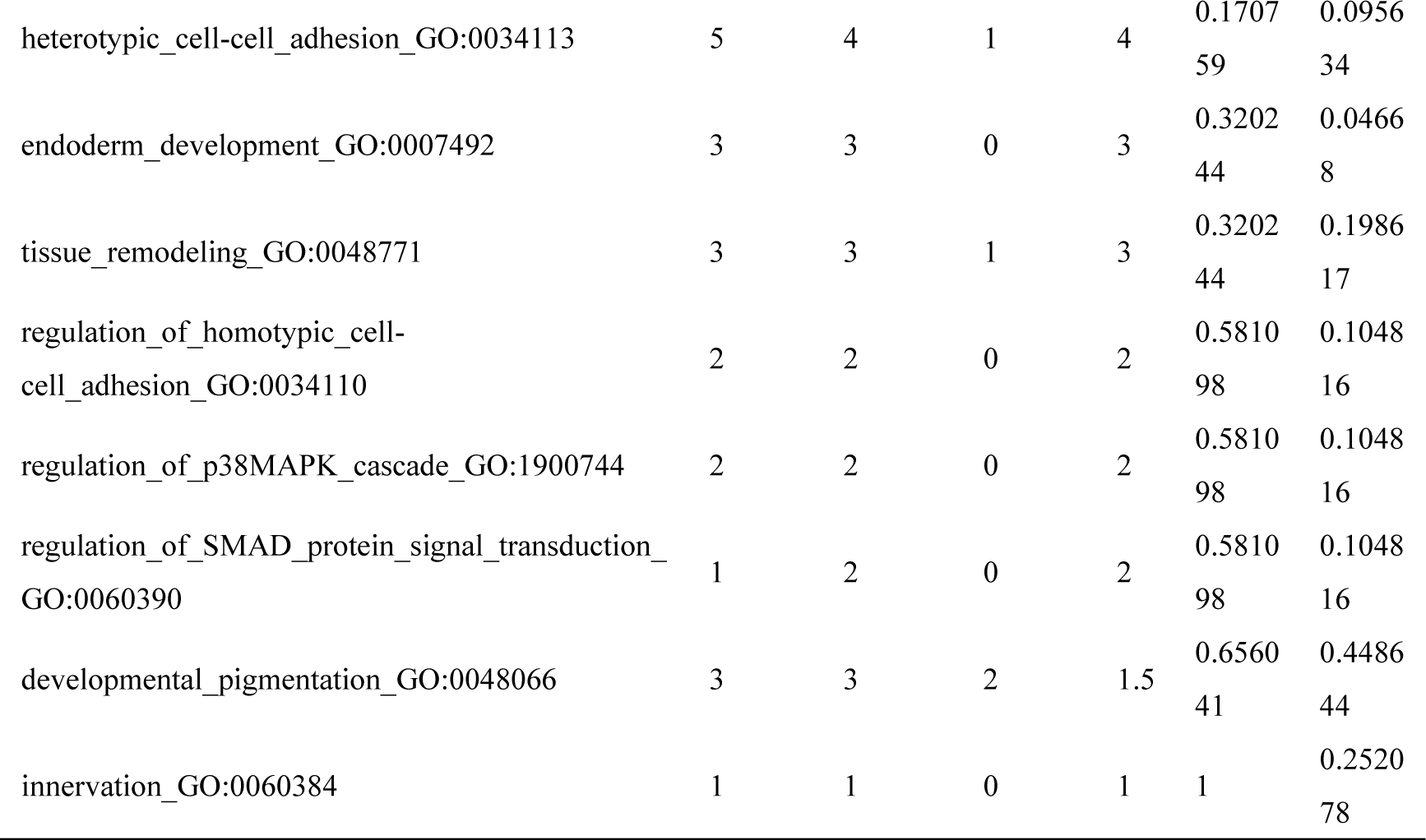
LoF excess in 21 biological processes enriched in the GO database

To assess the association between the 21 biological processes and the risk of CPT, the odd ratio (OR) is used to quantify the probability LoF mutations contributing to CPT. We defined the gene set including 5,173 genes from the 21 biological processes as the transcriptome high expression (THE). The gene set of biological process (GO:0008150) in GO includes 19,462 genes except THE, consisting of 14,254 genes, was referred to as the non-THE (NTHE). Comparing the OR of NTHE (1.73) between CPT cases and controls, a substantial increase to 18.29 in THE was displayed. This substantial rise in odds ratio demonstrates that the rare LoF excesses from THE significantly correlated with an increased risk of CPT risk. Ultimately, the specific genes contributing to CPT were identified as the those overlapping between the LoF-carrying genes and the THE. This resulted in the discovery of 151 CPT-contributing genes.

### Osteoclast differentiation with its regulated pathways contributes to CPT pathogenesis

To get insight into the molecular mechanism of these 151 CPT-contributing genes, they were queried in the Kyoto Encyclopedia of Genes and Genomes (KEGG) database for the enrichment that highlighted the pathway osteoclast differentiation (hsa04380, logP=-4.83) likely plays a potential role in CPT, along with its 3 regulated pathways including MAPK signaling (hsa04010, logP=-2.99), calcium signaling (hsa04020, logP=-2.95) and PI3K-Akt signaling (hsa04151, logP=-2.45). it was worth noting that tuberculosis pathway(hsa05152, logP=-4.59), Chemical carcinogenesis - DNA adducts pathway (hsa05204, logP=-4.56), as well as fluid shear stress and atherosclerosis pathway (hsa05418, logP=-4.57), were also identified(Fig3 a). To validate these findings, we examined the LoF excesses within these pathways. The results demonstrated significant LoF excesses in the MAPK signaling pathway (OR = 26, P = 1.45E-9), Tuberculosis pathway (OR = 12, P=1.37e-4), Chemical carcinogenesis-DNA adducts pathway (OR=10, P=0.001), PI3K-Akt signaling pathway (OR = 9, P = 1.48e-5), calcium signaling pathway (OR = 7, P = 2.66e-4), and osteoclast differentiation pathway (OR = 5.5, P = 0.001) in CPT cases (Fig3 b and Table 2).

**Fig3.**
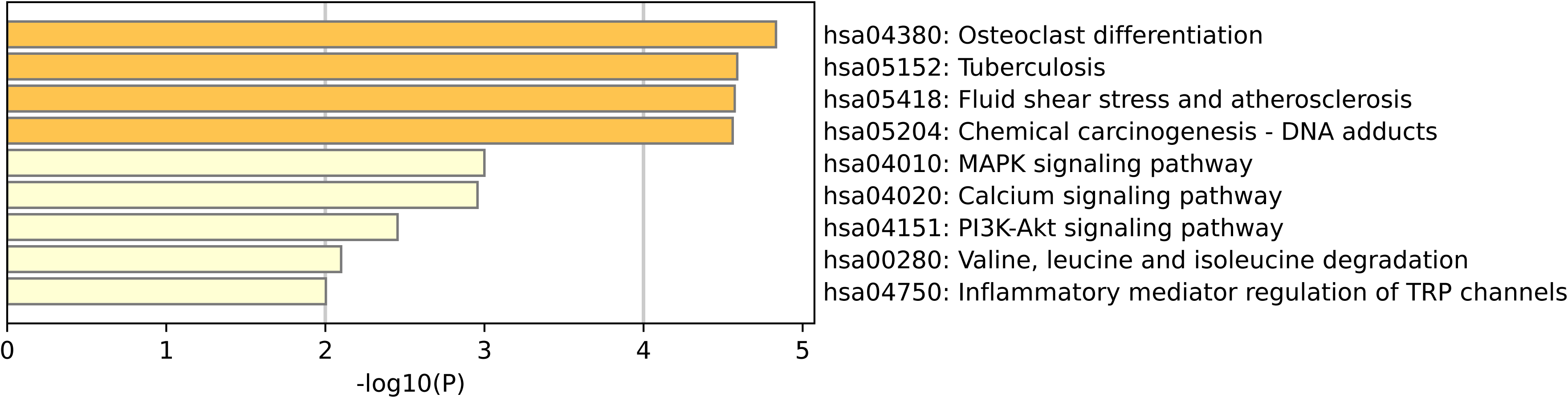
The KEGG pathway enrichment analysis and the LoF excesses of the transcriptome high expression (THE) gene set suffering from rare genomic LoF variations. (a) The KEGG enrichment of THE carrying the LoF mutations. X-axis indicates the -log(P) in which P is the probability that the genes are enriched in the pathway. Here the threshold of -log(P) is set as 2, indicating the P value 0.01. (b) the LoF excesses of the significant KEGG pathways enriched by THE. X-axis indicates the pathways from KEGG database and Y-axis indicates the odd ratio(OR) that the LoF mutations enrich in the pathway between 159 CPT cases and 208 controls. The numbers above the bars are the -log(P) in which P indicate the enrichment of LoF mutations in the pathway. Here the threshold of -log(P) is 2, indicating the threshold P 0.01.

**Table 2.** KEGG pathway enrichment of THE suffering from LoF variations

| Category | Term | Description | LogP | Log(q-value) | InTerm |
| --- | --- | --- | --- | --- | --- |
| KEGG Pathway | hsa04380 | Osteoclast differentiation | -4.83279 | -2.616 | 8/128 |
| KEGG Pathway | hsa05152 | Tuberculosis | -4.58891 | -2.616 | 9/180 |
| KEGG Pathway | hsa05418 | Fluid shear stress and atherosclerosis | -4.5734 | -2.616 | 8/139 |
| KEGG Pathway | hsa05204 | Chemical carcinogenesis - DNA adducts | -4.56047 | -2.616 | 6/69 |
| KEGG Pathway | hsa04010 | MAPK signaling pathway | -2.99947 | -1.453 | 9/294 |
| KEGG Pathway | hsa04020 | Calcium signaling pathway | -2.95648 | -1.451 | 8/240 |
| KEGG Pathway | hsa04151 | PI3K-Akt signaling pathway | -2.45406 | -1.112 | 9/354 |
| KEGG Pathway | hsa00280 | Valine, leucine and isoleucine degradation | -2.09919 | -0.836 | 3/48 |
| KEGG Pathway | hsa04750 | Inflammatory mediator regulation of TRP channels | -2.00287 | -0.799 | 4/98 |

Consistent with previous findings, the observed involvement in the osteoclast differentiation pathway and its 3 regulated pathways (MAPK signaling, calcium signaling and PI3K-Akt signaling) support the notion that CPT patients exhibit an imbalance in the low reparative potential of bone tissue due to the osteoclast activation prevailing over differentiation of osteoblasts, progenitor and mesenchymal cells [71]. The regulation of osteoclast differentiation and activation could become a promising strategy for preventing bone erosion in patients with CPT [17].

*NF1,* a reliable CPT-causing gene reported in previous studies, is believed to involve disruptions in bone remodeling and vascular abnormalities. Osteoclasts in *NF1* display enhanced resorption capacity and aberrant morphology [72]. The disorders of the Ras/MAPK pathway have an overlapping skeletal phenotype, such as scoliosis and osteopenia. The Ras proteins regulate cell proliferation and differentiation. Studies have shown that individuals carrying *NF1* mutations have osteoclast hyperactivity and increased bone resorption in the studies[73, 74]. In our study, *NF1* gene exhibits 9 LoF variations only in 12 patients (7.54%) with a heterozygous genotype. Alongside *AKT2*, *CD14*, *IL1RAP*, *MAP3K4*, *MAP3K11*, *RPS6KA1*, *TGFA* and *RASGRP2, NF1* was enriched in the MAPK signaling pathway (logP=-2.99). This suggests a potential framework for understanding how *NF1* contribute to the etiology of CPT.

Furthermore, *LILRB4*, *LILRB2*, *LILRB1* and *LILRA2* carrying 2 LoFs respectively are enriched in the osteoclast differentiation pathway along with *AKT2*, *CALCR*, *FCGR3A* and *IFNG*. The complex of *FCGR3A* and *LILRB1*/*LILRB2/LILRB4/LILRA2,* known as the osteoclast-associated receptor (OSCAR), is a regulator of osteoclast differentiation and dendritic cell maturation. It has been reported to cause skeleton-related phenotypes, such as abnormal osteoclast differentiation, reduced osteoarthritis manifestation, decreased chondrocyte apoptosis and abnormal bone morphology in knockout mice [75]. In osteoclasts, OSCAR is mediated by the immunoreceptor tyrosine-based activation motif adaptor protein FcRγ and co-stimulates osteoclasts cultured in an extracellular matrix or collagen [76]. *CALCR* is involved in regulating osteoclast-mediated bone resorption and has been reported to be associated with osteoporosis and bone mineral density quantitative trait locus 15 (https://www.genecards.org/). *IFNG* suppresses osteoclastogenesis by suppressing TRAF6-mediated signaling downstream of RANK, the receptor for RANKL [77]. That is, osteoclastogenesis would increase when the expression of *IFNG* is down-regulated due to the LoF variations.

Additionally, *COL6A5* carrying 2 LoF mutations, accompanied by *AKT2*, *JAK3*, *NOS3*, *TGFA*, *TLR2*, *SGK2*, *PKN3*, *TNN*, *KRT16* and *OPRM1*, is enriched in PI3K-Akt signaling pathway (logP = −2.45). This pathway has been reported to be involved in CPT by quantitative proteomics analysis [19]. The knockdown of *AKT2* was reported to suppress osteoclastogenesis through the PI3K/Akt/GSK3 signaling pathway [78]. Furthermore, alongside *HRC*, *MST1R*, *NOS3*, *P2RX1*, *TACR2*, *PPIF* and *TRDN*, *PLCZ1* carrying 2 LoF mutations is enriched in the calcium signaling pathway (logP = −2.95). Calcium signaling plays a crucial role in osteocyte mechanotransduction, as the release of calcium serves as a powerful second messenger. Calcium provides essential information for mechanical responses and participates in downstream regulatory processes [66].

Finally, *GSTM4* carrying 2 LoF mutations, along with *AKT2*, *IFNG*, *NOS3*, *PLAT*, *SELE*, *ARHGEF2*, *GSTO1*, *AKR1C4*, *AKR1C3*, *PPIF* and *SLC26A6*, is enriched in fluid shear stress and atherosclerosis (has05418). Previous studies have reported the involvement of osteoclasts and osteoblasts in activating different mechanical transduction pathways such as fluid shear stress and reported changings in their differentiation, formation, and functional mechanism induced by the application of different types of mechanical stress to bone tissue[79–81].

In summary, the osteoclast differentiation pathway, along with its 3 directly regulated pathways, not only exhibited rare LoF excesses but also showed shared transcriptome patterns between CPT cases and controls. These pathways serve as a reliable set of potential candidate pathways contributing to the etiology of CPT. Overall, there were 63 patients carrying 40 CPT-contributing genes with 56 LoF variations in the osteoclast differentiation pathways and their 3 directly regulated pathways (Table 3).

**Table 3.**
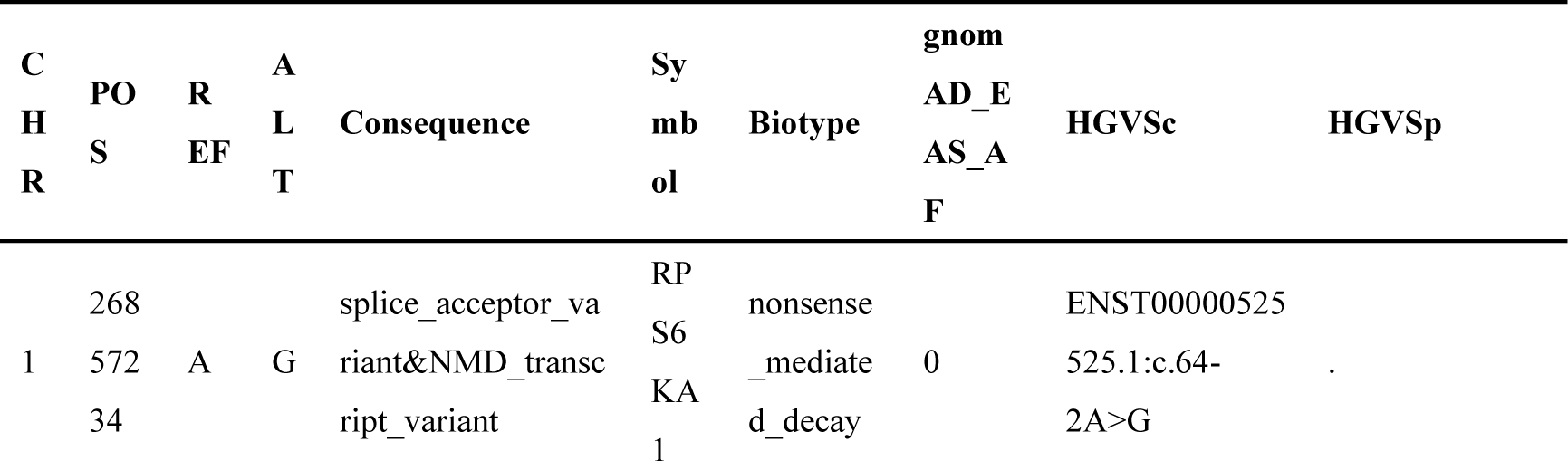
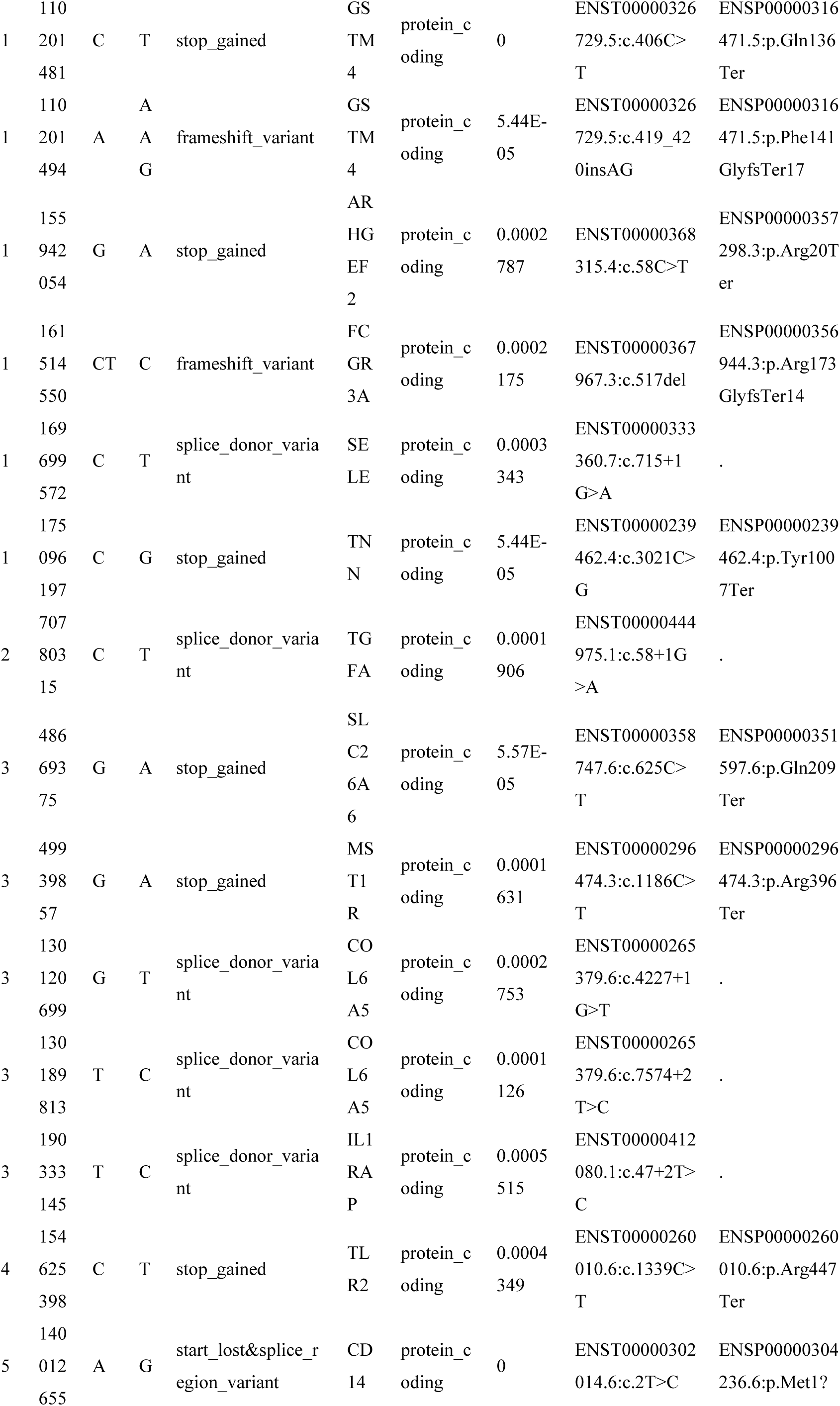
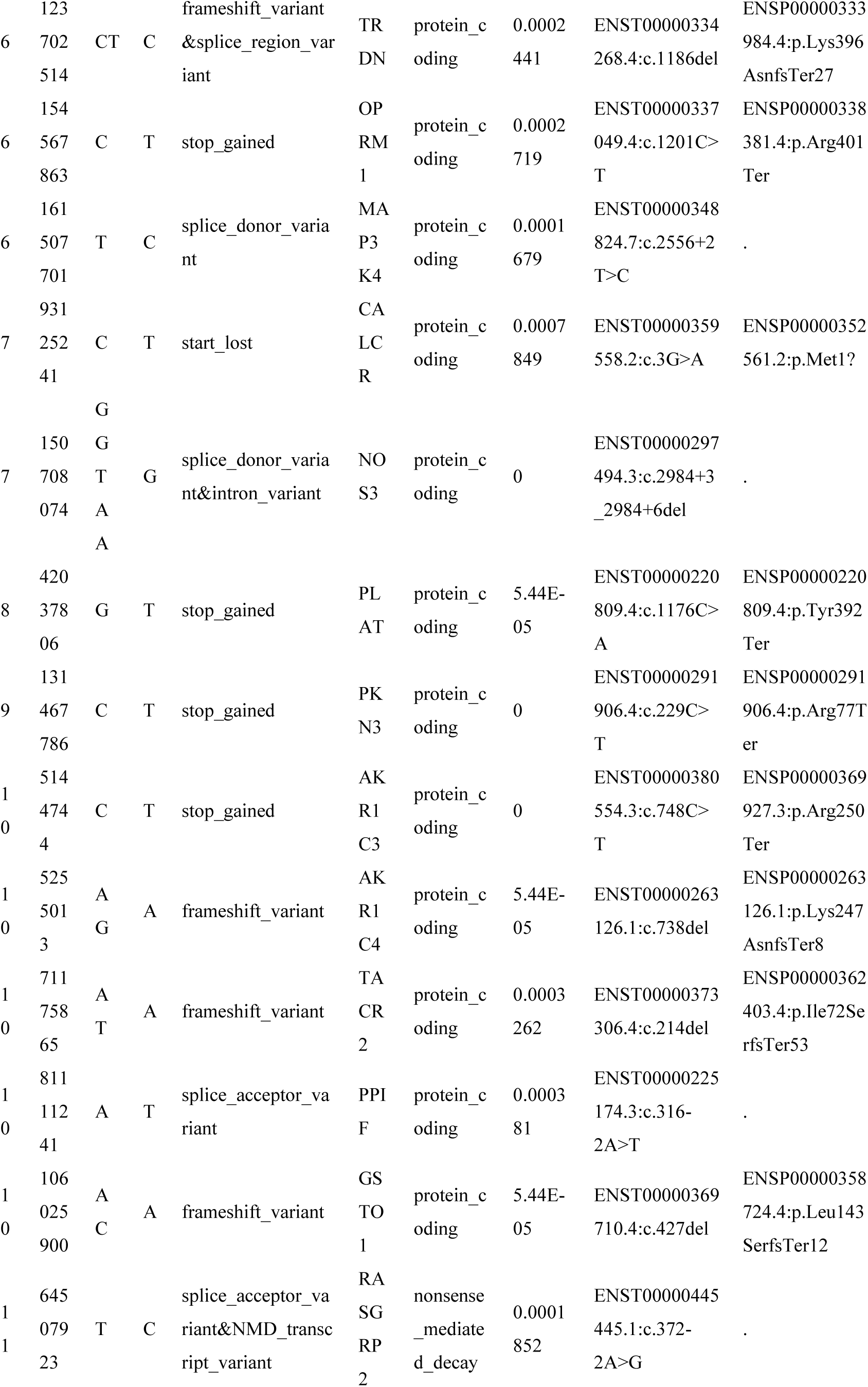
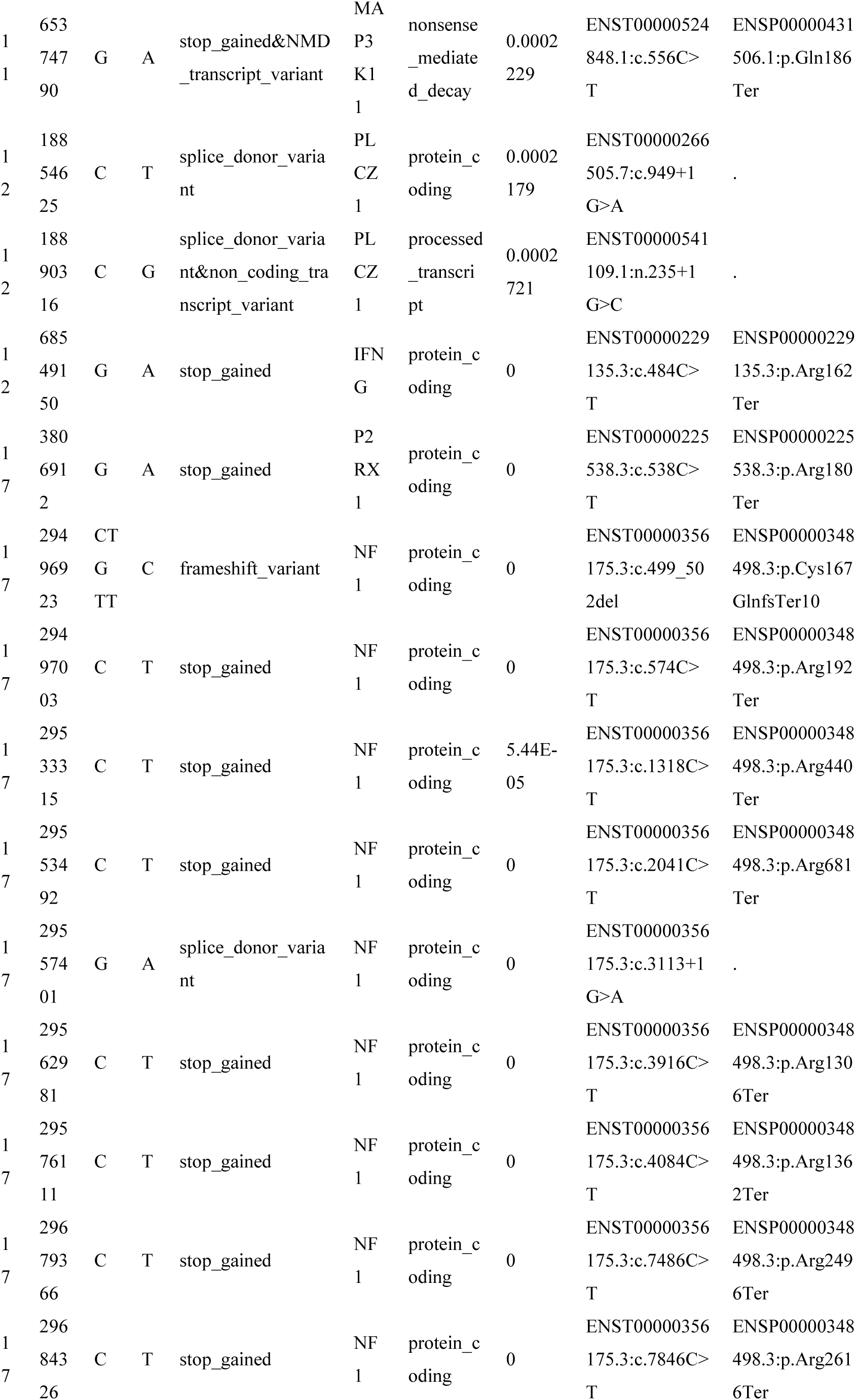
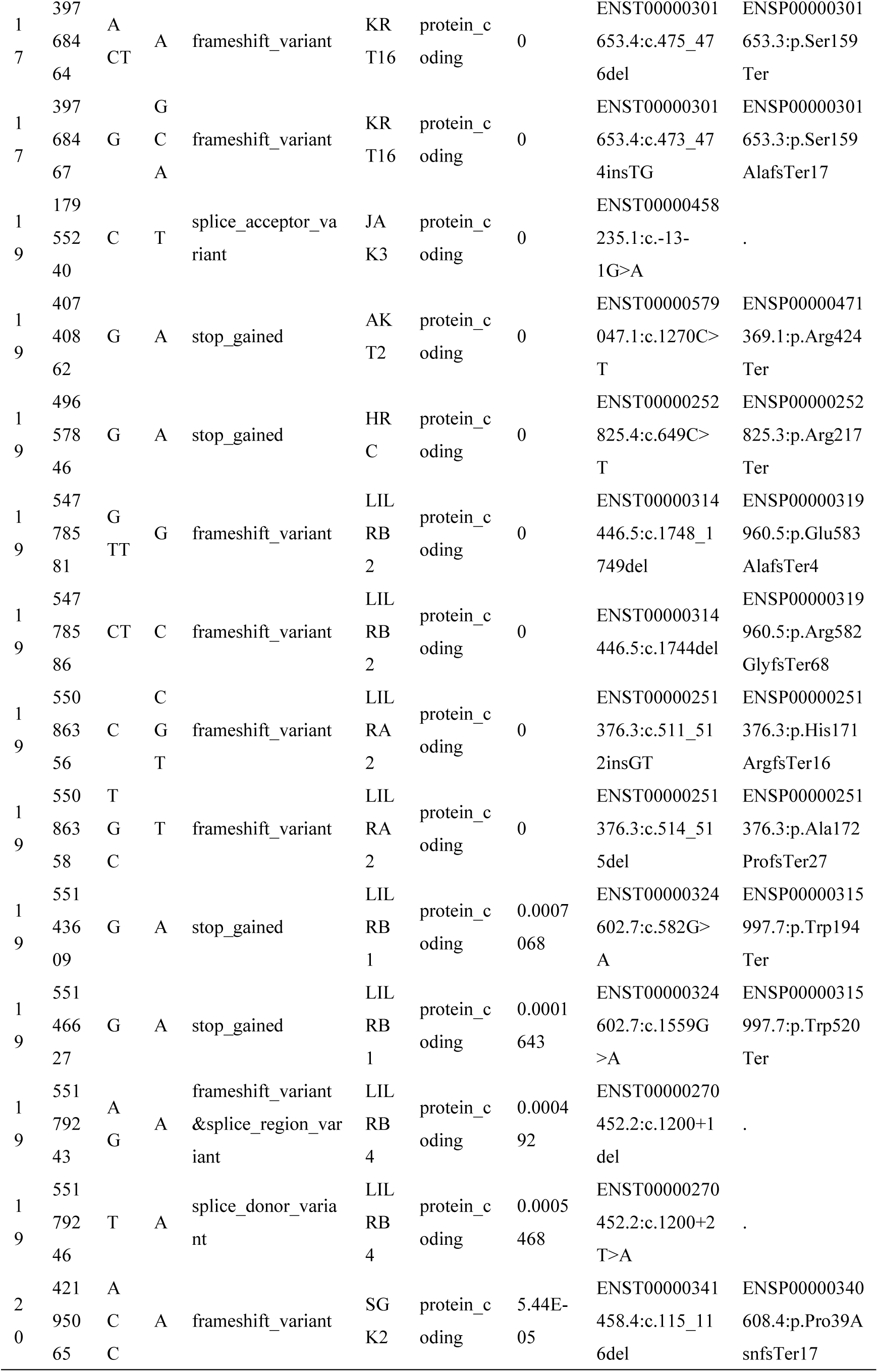
The list of CPT-causing LoF variations

### Hub genes enriched in hedgehog signaling pathway and likely regulate the molecular pathways of CPT

Interestingly, the Protein-protein interaction network analysis in the 151 CPT-contributing genes indicated that there were 8 hub genes (*BOC*, *SMO*, *PTCH2*, *ULK4*, *CORIN*, *USH2A*, *F12* and *MAGI3*) which showed a significant enrichment in hedgehog signaling pathway (hsa04340, logP= −6.5) and 3 genes *TIRAP*, *CD14* and *TLR2* in Lipid and atherosclerosis (hsa05417, logP=-6.5). As the members of Hedgehog signaling pathway, *BOC*, *SMO* and *PTCH2* respectively carried 1 LoF mutation, and was indicated to be regulated by the other 5 hub genes in the PPI network. Hedgehog signaling pathway had been reported play a crucial role in regulating bone formation and homeostasis[82, 83] and the premature SHH signaling resulting from disruption of *GPR161* causes defects in limb and skeletal morphogenesis[84].

## Discussion

CPT is a rare bone disorder primarily affecting newborns and characterized by challenging complications such as refracture, tibial non-union, and failed surgery [85]. The underlying pathophysiology involves excessive osteoclast activity caused by the fibrous hamartoma surrounding the bone and impaired osteogenesis and bone morphogenetic protein function, resulting in frequent tibial fractures, bone atrophy, and compromised bone remodeling [86]. Neurofibromatosis type 1, with the prevalence of 1/3,000 [87], is linked to CPT in many cases and contribute to anterolateral bowing of the tibia, leading to pathologic fracture. Previous study has claimed the gene *NF1* encodes neurofibromin, a ubiquitous protein expressed in osteoblasts, osteoclasts, chondrocytes, fibroblasts and endothelial cells, and neurofibromin negatively regulates the activity of Ras which involved in cellular proliferation and differentiation, leading to the physically preventing bony bridging and endochondral bone repairing by the recurrent invasion and increased proliferation of fibrous lesion cells in the cellular deficiencies, suggesting the role of *NF1* in bone development, homeostatic regulation, and repair in Neurofibromatosis type 1 [88]. However, the complete understanding of CPT cannot be fully attributed to the *NF1* gene within the framework of neurofibromatosis type 1. And other factors, including intrauterine trauma and generalized metabolic disturbances at the time of birth, are believed to contribute [89]. Recent studies have identified several biological processes, such as skeletal system development, tissue morphogenesis and cell differentiation/proliferation, are associated to CPT[18, 19, 53], these findings suggest the involvement of intricate molecular processes in which other genes, in cooperation with *NF1*, may contribute to CPT. However, the precise mechanisms underlying these effects have not yet been elucidated.

In this study, we sequenced the transcriptome of periosteal tissues from three patients with CPT and two healthy individuals, and identified 180 up-regulated genes in CPT patients, which were enriched in 40 biological processes related to bone homeostasis. These biological processes encompassed skeletal system development, tissue remodeling (e.g., bone resorption), cell matrix assembly, regulation of cell adhesion, regulation of the MAPK cascade and autophagy in bone metabolism. These findings are consistent with previous studies and provide an overview of the biological processes likely associated with CPT, despite the limited number of patient samples included in this analysis. Furthermore, to gain insight into the effects of genomic LoF variations on CPT, we performed WES on 159 patients and observed significant excesses of rare LoF variations in 21 out of the 40 biological processes. This indicates the rare LoF variations play an important role in CPT-related biological processes. Most importantly, through gene-pathway enrichment analysis using the KEGG database, we identified 63 patients carrying 40 genes with 56 LoF variations in the osteoclast differentiation pathway with its 3 directly regulated pathways (MAPK signaling pathway, Calcium signaling pathway and PI3K-Akt signaling pathway), as well as the fluid shear stress and atherosclerosis pathway. This provides a comprehensive illustration of how rare genomic LoF variations in the osteoclast differentiation pathways contribute to the pathological process of CPT.

Additionally, we also observed that 19 out of the 40 biological processes showed significant gene enrichment in the transcriptome but did not exhibit significant LoF excesses. This suggests the involvement of other types of genomic mutations contributing to CPT. For instance, among the CPT patients, apart from MAPK signaling pathway(P=3.67e-08), calcium signaling pathway (P=1.58e-10) and PI3K-Akt signaling pathway (P=4.15e-10), we found missense mutations exhibited the excesses specifically in apoptosis pathways (P=6.58e-05) and mTOR signaling pathway (P=3.15e-05) which have been confirmed as the crucial pathways orchestrating the bone resorption in bone metabolic diseases [90, 91].

Compared with the fact that up to 80%-84% of patients with CPT present with neurofibromatosis type 1, we found NF1 carried 9 LoF mutations in 12 patients, with a prevalence 19% in this study. This paradox in prevalence suggests some of our 56 LoF mutations may not directly cause CPT but instead participated in the regulation of the imbalanced bone homeostasis resulted from CPT. Furtherly considering the extreme low prevalence of CPT, de novo mutations would serve as more accurate genomic markers for the genetic diagnosis of CPT under our current pathway framework.

In summary, our findings highlight that rare genomic LoF variations in genes involved in osteoclast differentiation and regulation pathways play crucial role in the pathological process of CPT. This provides a new framework for understanding how genetic variations regulate biological processes in the pathology of CPT.

## Method

### Patient Recruitment and study subjects

A consecutive cohort of 165 CPT patients admitted into the department of Orthopedics of Hunan Children’s Hospital from 2016 to 2020 was enrolled in this study. For WES, a total of 159 CPT patients, of which 75 cases had been reported in 2019[7], provided peripheral blood samples for DNA extraction and sequencing. For the transcriptome sequencing, two normal periosteal tissues were obtained from healthy children undergoing surgery for tibial fracture, as well as three diseased periosteum tissues were obtained from the patients with CPT during surgery at Hunan Children’s Hospital. This study had been approved by the Ethics Committee of Hunan Children’s Hospital, and informed consent had been obtained from all patients.

### Transcriptome sequencing by nanopore platform and data analysis

#### RNA Extraction from Periosteal Tissue

During the surgical procedure, periosteal tissue devoid of bone needles and encompassing cortical bone was collected and immediately dissected into small fragments measuring approximately 200 mg with a diameter of approximately 5 mm. These fragments were then placed in centrifuge tubes filled with phosphate-buffered saline (PBS) and thoroughly washed. Subsequently, they were transferred to cryovials and snap-frozen in liquid nitrogen for storage. Total RNA extraction from the periosteal tissue was performed using Takara TRIzol reagent according to the manufacturer’s standard protocol. The extracted RNA was subjected to quality assessment using the IMPLEN Nano Photometer for spectrophotometric analysis, Nanodrop 2000 for concentration measurement of RNA samples, Agilent 2100 for integrity assessment, and agarose gel electrophoresis. One microgram of purified total RNA with a minimum RNA integrity number (RIN) value of 6 was reverse transcribed into complementary DNA (cDNA) using the cDNA-PCR sequencing kit (SQK-PCS109).

#### Full-Length Transcriptome Sequencing

The experimental workflow followed the standard procedure provided by Oxford Nanopore Technologies (ONT), including sample quality check, library construction, library quality assessment, and library sequencing. The total RNA extracted from periosteal tissue was required to be at least 1.5 μg, suitable for three library preparations, with a concentration of ≥40 ng/μl, a volume of ≥10 μl, an OD260/280 ratio between 1.7 and 2.5, an OD260/230 ratio between 0.5 and 2.5, and a normal peak at 260 nm absorption. Samples with an RIN value of 6 or higher were selected for full-length transcriptome sequencing using ONT. The library construction process involved the following steps: First, oligo dT primers were utilized to anneal with the polyA tail of mRNA, followed by the addition of reverse transcriptase (RT) to synthesize the first cDNA strand. Next, specific PCR adapters were added to both ends of the first cDNA strand, and a 14-cycle cDNA PCR was performed using LongAmp Tag DNA polymerase (New England Biolabs, Ipswich) with an extension time of 8 minutes. This step amplified the double-stranded cDNA using amplification primers. Subsequently, DNA damage repair and end repair were conducted using the NEBNext FFPE DNA Repair Mix and NEBNext Ultra II End Repair/dA-Tailing Module to repair and end-repair the nucleic acid fragments, followed by A-tailing. The sequencing adapters were then ligated using the SQK-LSK110 kit (ONT), and purification was carried out using Agencourt XP beads (Beckman Coulter, USA). The prepared cDNA library was sequenced using the Oxford Nanopore PromethION 48 platform (Biomarker, Beijing).

After transcriptome sequencing, we trim the adaptors by software porechop v0.2.4(https://github.com/rrwick/Porechop), aligned the reads to reference hg19 and quantified gene expression using the FLAIR software [92]. We identified 179 significantly up-regulated genes and 134 significantly down-regulated genes (|logFC| > 2 and FDR < 0.01) between CPT cases and controls using the differential expression analysis module of the Trinity software [93]. Furthermore, we obtained the significantly enriched pathways (supplementary table 1) of these 179 up-regulated genes using the Metascape software [94] (accessed May 2022).

### WES, variation calling and filtering

Genomic DNA from peripheral blood was extracted using the standard phenol-chloroform method. The DNA of all 159 CPT patients was fragmented and the exome was captured using the Agilent SureSelect Human All Exon V6 kit. The captured DNA was sequenced with 100 bp pair-end reads using the BGISEQ2000 platform, following the manufacturer’s instructions. Each sample yielded an average of 13.3 Gb of raw data with an average depth of 132 and a coverage ratio of 99.84% of the captured target region of 60.46 M. Over 98% (average ∼92.9%) of the bases had a phred quality score > 30. For the variation calling, the raw reads were filtered by SOAPnuke [95] and then mapped to the human genome reference (UCSC/GRCh37/hg19) using the Burrows-Wheeler Aligner (BWA-MEM, version 0.7.10) [96]. Variants were called using the Genome Analysis Tool Kit (GATK, version 4.1) [97]. To validate the accuracy of the variation calling, 208 Chinese Han individuals from the 1000 Genomes Project were selected as a control to perform principal component analysis (PCA). This allowed us to remove the false positive WES variations by setting genotype quality > 30, the minimum read depth of 20 for each genotype, and an additional setting of the allele balance of 0.3 for each heterozygous genotype. Furthermore, all indels with allele lengths greater than 10 bp were removed. The Variant Effect Predictor (VEP) [98] was used to annotate and classify all the variants. Subsequently, the variants were filtered based on their frequency in public database gnomAD (https://gnomad.broadinstitute.org/) and the bi-allelic LoF variants (frameshift_variant, splice_acceptor_variant, splice_donor_variant, start_lost, stop_gained and stop_lost) with MAF < 0.001 were retained.

### LoF excess test for pathway gene sets

After obtaining the VCF file of LoF variations, the software rvtest[99] was used to test the LoF excess for each gene set from the biological processes or KEGG pathways. The burden test was performed using the CMC model, with a control population consisting of 208 Chinese Han individuals selected from the 1000 Genomes Project as the control population against the 159 patients with CPT. The VCF files containing the variations information of both the 159 patients and the 208 control individuals were at the service for request. The genes corresponding to the biological processes were obtained from http://amigo.geneontology.org/amigo/term/GO:0008150, with the organism selected as Homo sapiens. The genes of KEGG pathways were downloaded from https://www.genome.jp/kegg/.

## Data availability

Both RNA-seq and WES data are at the service for request.

## Authors information

Hui Huang, Huanhuan Peng and Haibo Mei conceived and designed the study. Guanghui Zhu and Yu Zheng collected the samples and performed the experimental analysis. Nan Li performed the data analysis and wrote the manuscript. All the authors read and approved the final manuscript.

## Ethics declarations

The experimental protocol was approved by the Research Ethics Committee of Hunan Children’s Hospital. Informed consent was obtained from every patient.

## Data Availability

All data produced in the present study are available upon reasonable request to the authors

https://pan.genomics.cn/ucdisk/s/N73UJ3

---

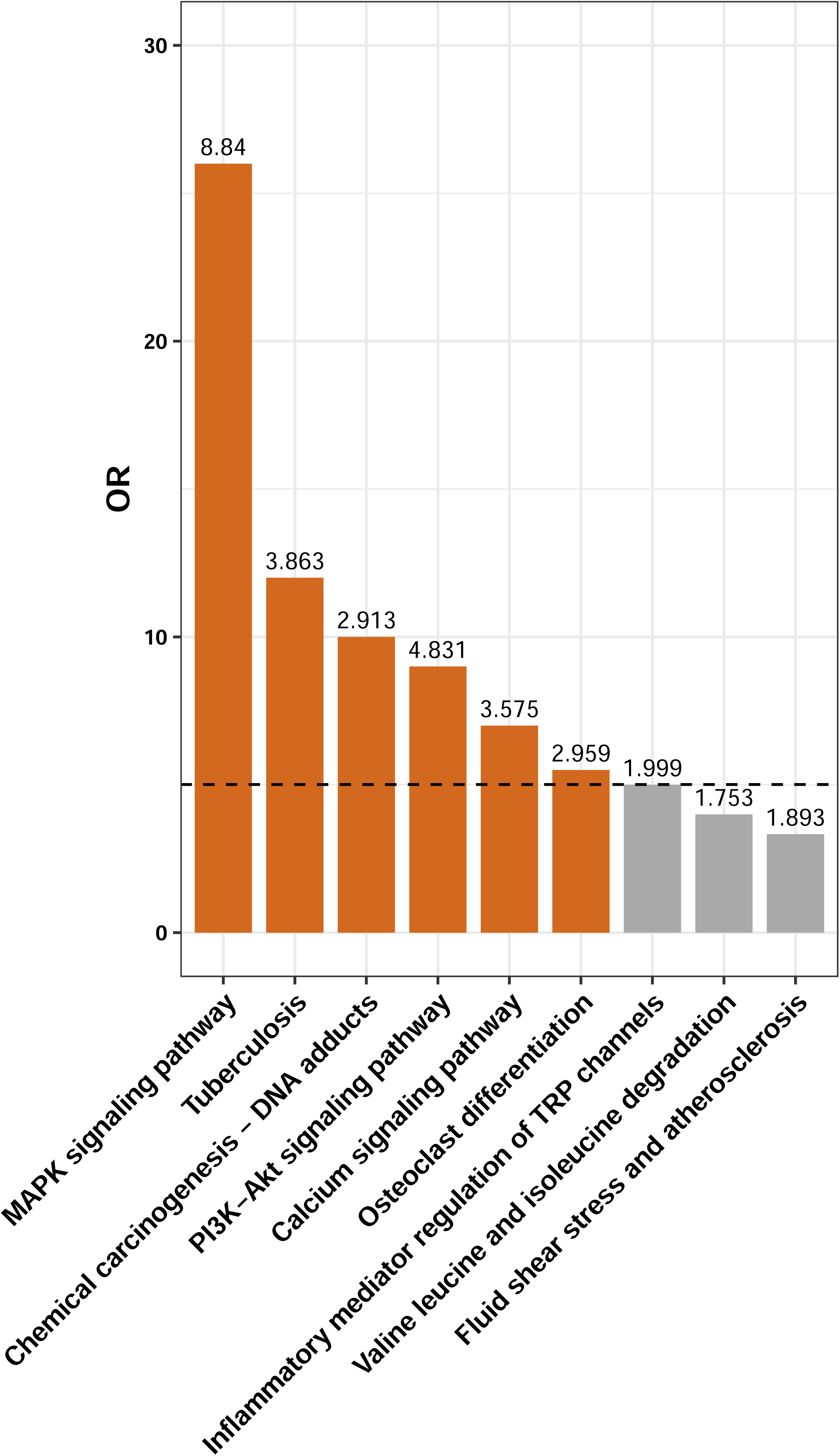

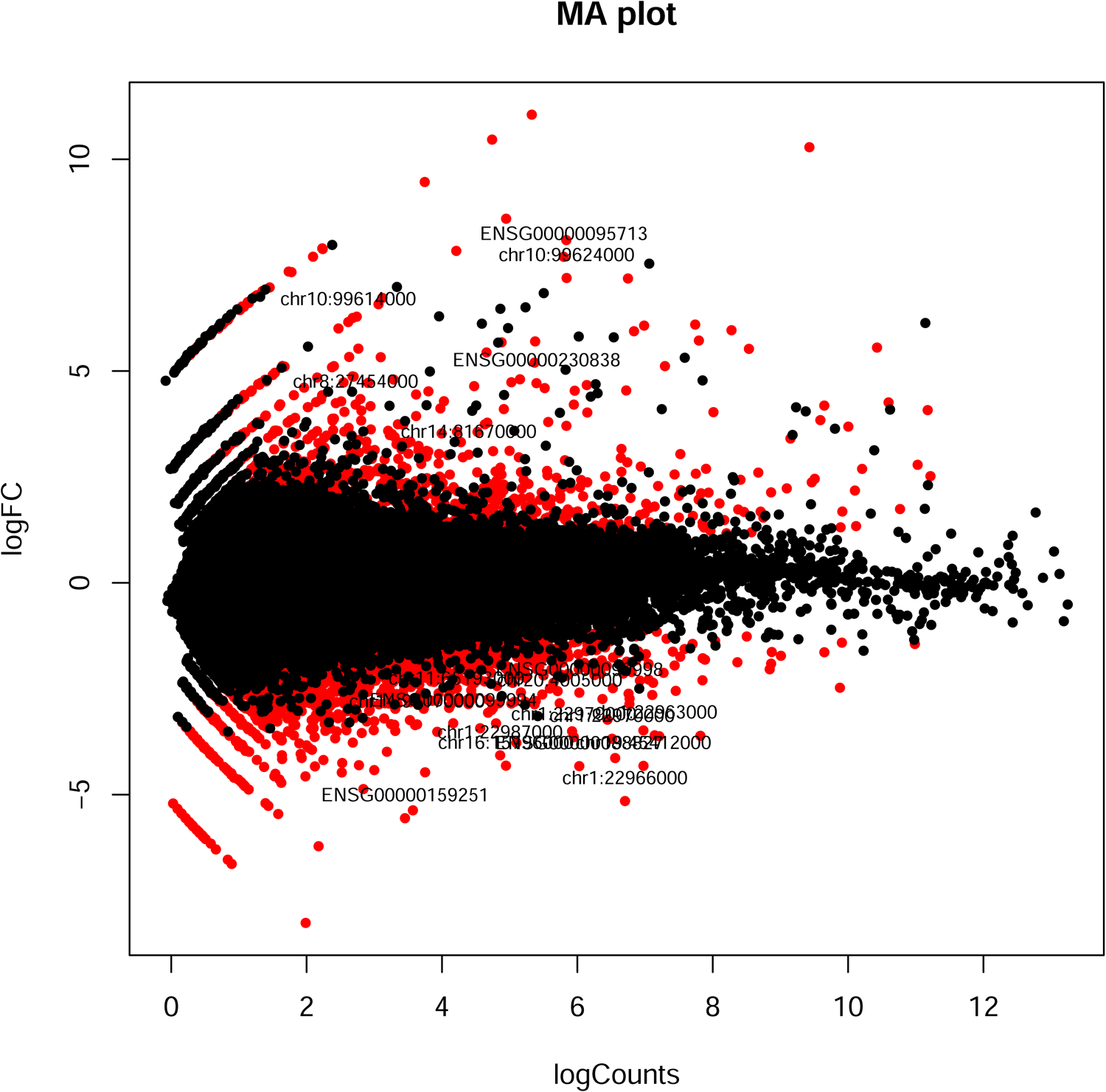

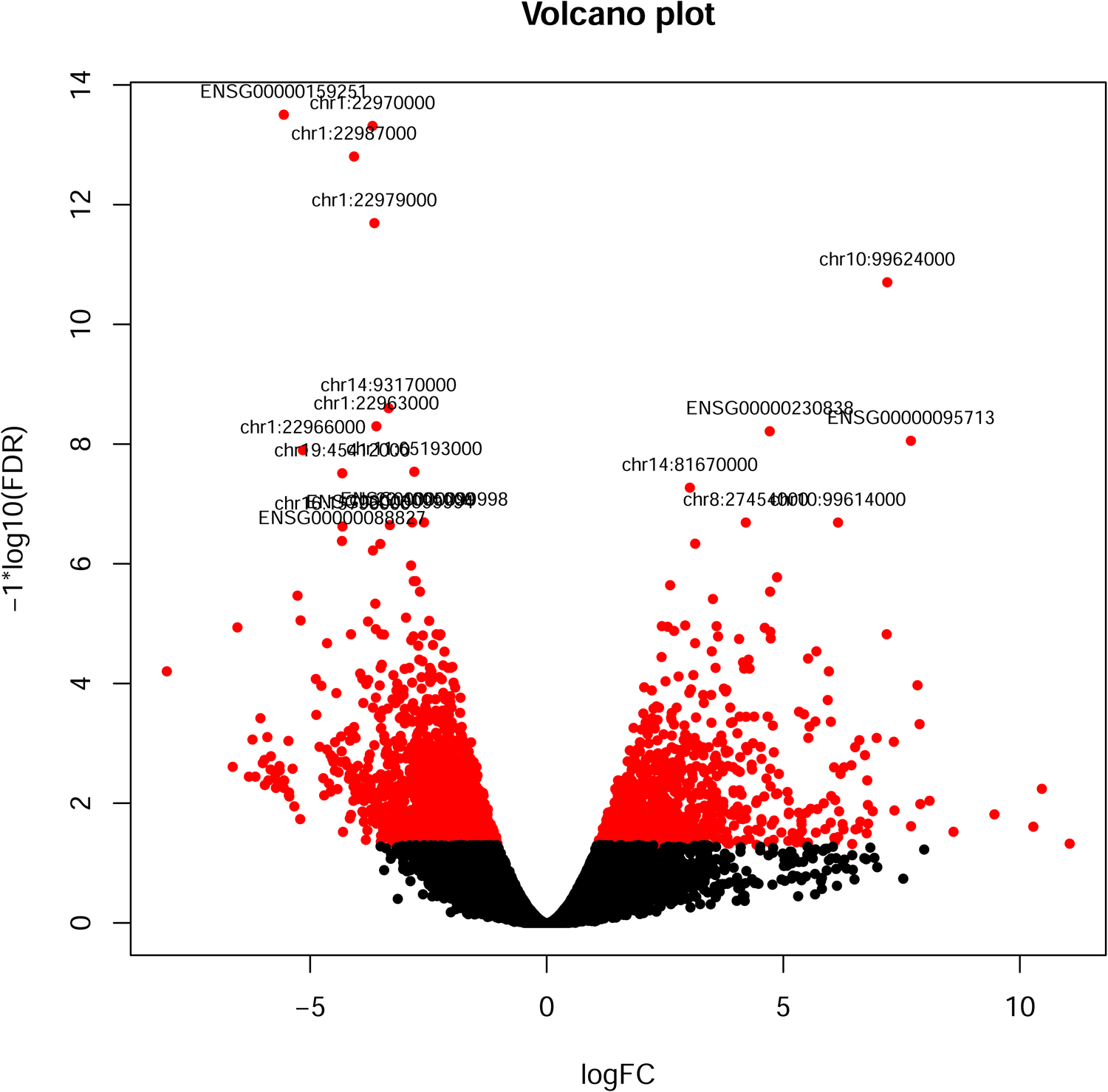

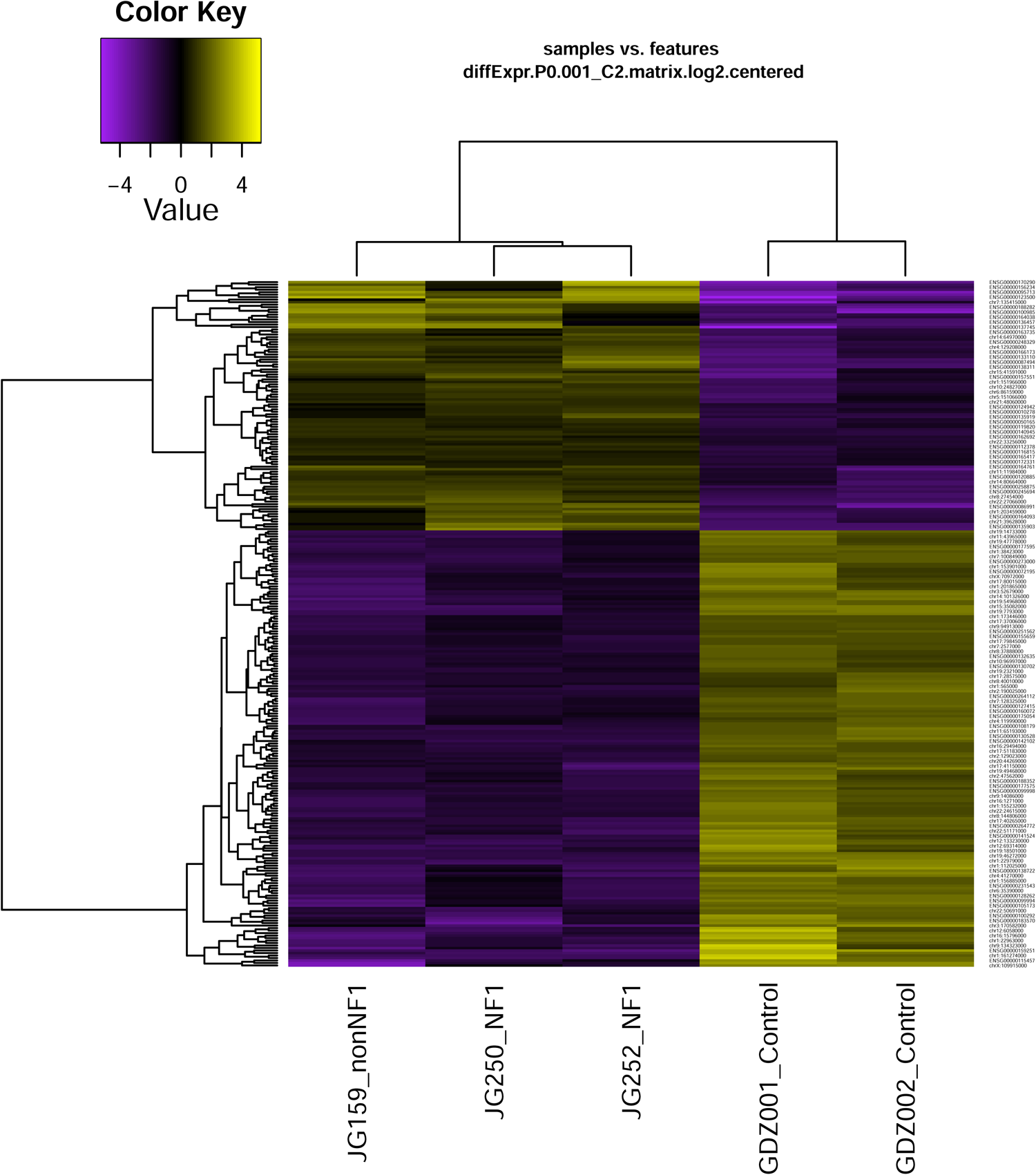

