## Supplementary figures and images for "Unraveling the Molecular Landscape of Congenital Pseudoarthrosis of the Tibia: Insights from a Comprehensive Analysis of 162 Probands"

### Supplementary Fig1

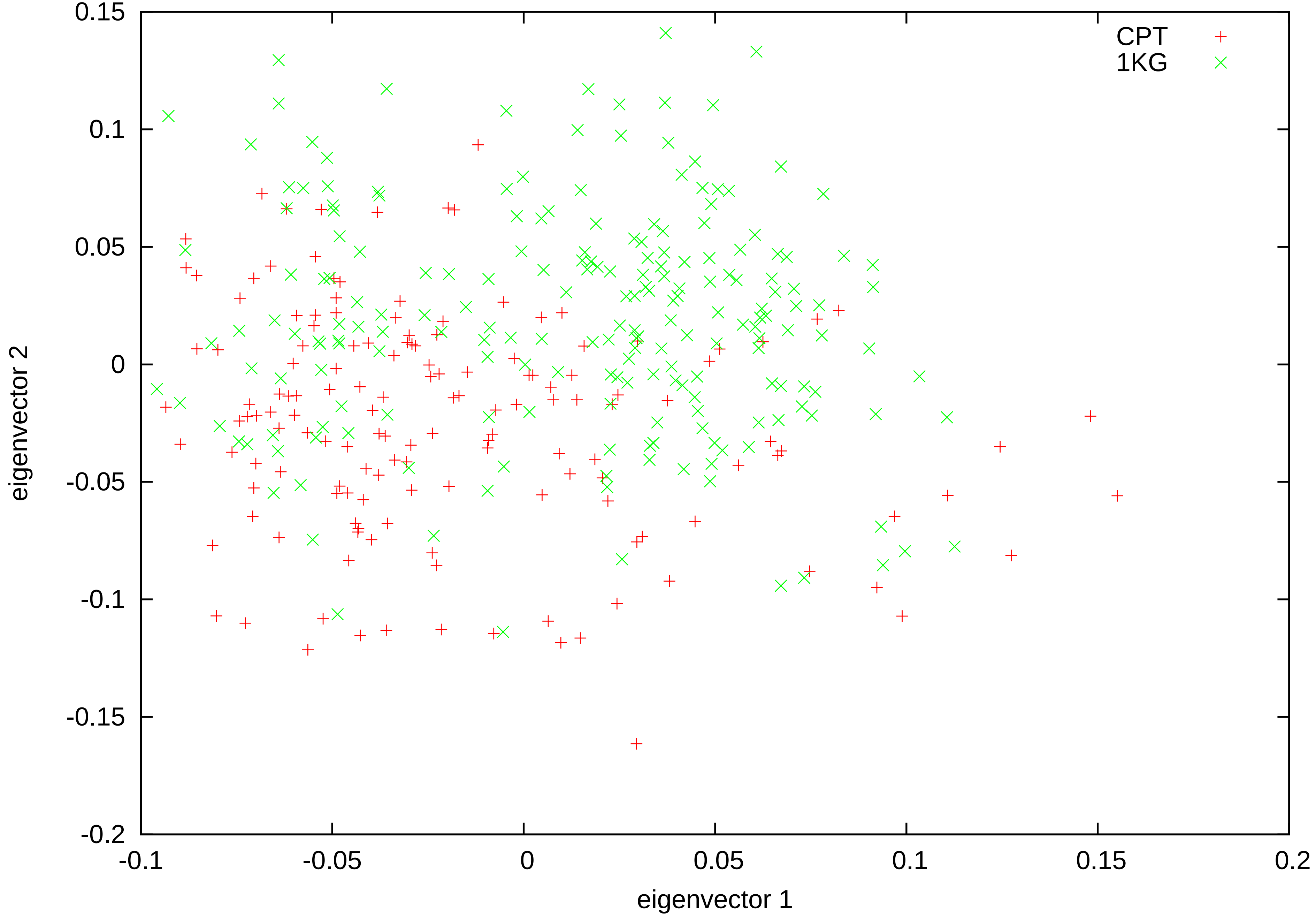
